# The Cardiac Efficiency Index: A Novel Parameter to assess Left Ventricular Diastolic Performance

**DOI:** 10.64898/2026.11.24.26368919

**Authors:** Jeevan Rajkumar, Sonakshi Manjunath, Majesh Makan, Jonathan Moreno, John Davis, Adam May

## Abstract

**BACKGROUND:** Standard echocardiographic metrics of diastolic function assess ventricular relaxation and filling pressures largely independent of systolic performance.

**OBJECTIVES:** To introduce and evaluate the Cardiac Efficiency Index (CEI), a novel echocardiographic parameter integrating mitral inflow kinetics with forward stroke volume to provide a more integrated assessment of diastolic physiology.

**METHODS:** We convenience sampled 82 patients with recent transthoracic echocardiography. CEI was evaluated against American Society of Echocardiography (ASE) diastolic dysfunction (DD) grade and left atrial pressure (LAP; normal or elevated). Binary logistic regression evaluated discrimination of elevated LAP and ASE grade ≥2. Ordinal logistic regression evaluated increasing ASE grade (0–1, 2, 3).

**RESULTS:** Elevated LAP was present in 33/80 (41.3%) patients with valid assessment while 22/66 (33.3%) patients with valid ASE grading had grade ≥2 DD. For elevated LAP, CEI yielded an AUROC of 0.954 [95% confidence interval (CI), 0.898–0.993] compared with 0.949 [0.895–0.988] for E/e′. For ASE grade ≥2 AUROC was 0.941 [0.854–0.994] for CEI and 0.942 [0.868–0.991] for E/e′. Ordinal regression demonstrated comparable discrimination (c-index 0.904 [CEI] vs. 0.912 [E/e′]) while CEI had a larger per-SD association with increasing ASE grade (OR 10.45 [4.03–27.08]) than E/e′ (OR 7.58 [2.92–19.67]). In the combined model, CEI remained independently associated with increasing ASE grade (OR 6.82 [2.24–20.73]; P<0.001) whereas E/e′ did not (OR 1.75 [0.70–4.38]; P=0.23).

**CONCLUSIONS:** CEI provided strong discrimination of elevated LAP and ASE DD grade in this proof-of-concept analysis, supporting further validation against invasive hemodynamics in larger cohorts.

## Introduction

Traditionally, echocardiographic assessment of diastolic function has focused on characterizing ventricular relaxation and estimating filling pressures using parameters derived from transmitral flow, mitral annular motion, and pulmonary venous flow.^1,2^ Contemporary ASE guidelines reflect the complexity of diastolic assessment by integrating multiple echocardiographic parameters to estimate left ventricular filling pressures and classify diastolic dysfunction.^2^ However, individual parameters are influenced by loading conditions, cardiac structure, rhythm, and other physiologic factors, and discordance among individual measurements and reference standards remains common.^3–7^ Although a multiparametric approach helps address the limitations of individual indices, conventional methods largely evaluate diastolic physiology independently of the forward stroke volume (SV) generated from ventricular filling. Yet, ventricular filling and forward output are intrinsically linked, as systolic performance depends in part on the diastolic properties governing ventricular filling.

Transmitral flow, measured by pulsed-wave (PW) Doppler, reflects the integrated effects of ventricular relaxation, chamber compliance, preload, left atrial function, and heart rate. As the principal route of left ventricular inflow, coupling early transmitral inflow kinetics during diastole to the resulting forward SV may provide physiologic information not captured by conventional assessments of diastolic function or filling pressures alone.

These considerations highlight the need for complementary approaches that characterize diastolic physiology in relation to forward cardiac output. By integrating early mitral inflow kinetics with forward stroke volume (SV), we introduce the Cardiac Efficiency Index (CEI), a novel echocardiographic parameter designed to relate the hemodynamic characteristics of ventricular filling to the forward output generated. In this paper, we first describe the conceptual basis and derivation of CEI and then perform a proof-of-concept retrospective cohort analysis to evaluate its ability to discriminate left atrial pressure (LAP) and ASE diastolic dysfunction grade.

## Methods

### Cardiac Efficiency Index

#### Early Diastolic Inflow Kinetics

Early diastolic filling begins with mitral valve opening and is governed by the evolving pressure gradient between the left atrium (LA) and left ventricle (LV). Foundational combined Doppler-hemodynamic studies established that transmitral flow patterns reflect the interaction between ventricular relaxation and loading conditions, with rising LAP augmenting early diastolic filling despite impaired LV relaxation.^8^ On pulsed-wave (PW) Doppler echocardiography, E-wave velocity (E) reflects the peak velocity of early diastolic filling. Peak E-wave acceleration time (PEWAT) is the interval from the onset of transmitral flow to peak E-wave velocity, providing complementary information regarding the temporal dynamics of early diastolic inflow. ^9^ These two Doppler-derived parameters (Error! Reference source not found.) characterize the initial rapid-filling period, during which a substantial proportion of ventricular filling occurs. In a variety of pathophysiologic states in which decompensated hearts have acutely elevated filling pressures, a greater proportion of ventricular filling occurs during this early diastolic phase.^10–12^ Consistent with this physiology, prior work examining E-wave acceleration demonstrated progressively shorter onset-to-peak E-wave acceleration times and higher acceleration rates with advancing diastolic dysfunction.^13^

Using the modified Bernoulli equation (Δ**P = 4v²**) to relate E-wave velocity to the instantaneous transmitral pressure gradient, the rate of LA-to-LV pressure-gradient development can be expressed as:

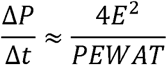

where P is pressure, t is time, E is the peak E-wave velocity (m/s), and PEWAT is the time (ms) from E-wave onset to peak velocity. Thus, *4E²/PEWAT* serves as the numerator of CEI, integrating the magnitude of the estimated transmitral pressure gradient with the interval over which it develops during early diastole.

#### Normalization to Forward Stroke Volume

Early mitral inflow kinetics help describe the characteristics of ventricular filling but not the forward output ultimately generated from that filling. In the absence of significant valvular regurgitation or intracardiac shunting, diastolic LV inflow is reflected in systolic LV outflow. Stroke volume (SV), measured using left ventricular outflow tract (LVOT) PW Doppler (via the product of LVOT velocity-time intergral [VTI] and LVOT cross-sectional area), therefore provides a physiologically complementary measure of the forward output generated from ventricular filling. Accordingly, SV indexed to body surface area (SVI) was incorporated as the denominator of CEI, relating early mitral inflow kinetics to the resulting indexed forward SV:

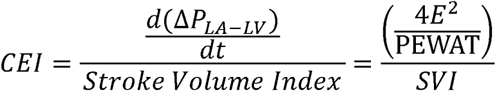

#### Physiologic Interpretation of the Cardiac Efficiency Index

The relationship between mitral inflow kinetics and forward SV provides the conceptual basis for the Cardiac Efficiency Index (CEI). Rather than evaluating early diastolic pressure-gradient development in isolation, CEI relates the hemodynamic characteristics of diastolic filling to the forward output generated.

**Figure 2** illustrates the physiologic basis of CEI using pressure-volume loops representing a normal heart and a decompensated heart with reduced ejection fraction. In the diseased state, the ventricle operates at a higher and steeper portion of the end-diastolic pressure-volume relationship (EDPVR), where incremental increases in preload are associated with greater increases in ventricular pressure and reduced forward SV. This physiology is reflected by greater LA-to-LV pressure-gradient development during early diastole (higher *4E*^2^*/PEWAT*) coupled with reduced SVI, resulting in a high CEI. Conversely, the normal ventricle operates along a less steep portion of the EDPVR, where incremental increases in preload are associated with smaller changes in ventricular pressure while preserving a relatively larger forward SVI.

The combination of lower *4E*^2^*/PEWAT* and greater SVI therefore results in a low CEI. These contrasting physiologic states illustrate how CEI relates diastolic filling hemodynamics to the efficiency of forward flow generation. Representative clinical examples across different physiologic states are provided in **Supplementary Table S1.**

In this framework, higher CEI values reflect greater early mitral inflow kinetics relative to the forward SVI generated, consistent with less efficient translation of ventricular filling into forward output. Conversely, lower CEI values reflect greater forward SVI relative to early mitral inflow kinetics, consistent with more efficient translation of ventricular filling into forward output.

#### Study design and population

To evaluate the feasibility and potential utility of this novel parameter, we conducted a retrospective proof-of-concept cohort study using echocardiographic data obtained across the BJC HealthCare system, including Barnes-Jewish Hospital, Missouri Baptist Medical Center, Christian Hospital, and affiliated hospitals, through an integrated echocardiography platform. The Washington University’s Institutional Review Board approved the study and its design. De-identified data and scripts for statistical analyses are available on request. Adults aged 18 years or older, either inpatient or outpatient, undergoing transthoracic echocardiography (TTE) for any clinical indication were eligible. To ensure distribution across the spectrum of diastolic physiology, patients who had undergone echocardiography within a 60-day window at study initiation were convenience sampled across institutionally derived tertiles of E/e′ (< 8, 8–12, > 12). We sequentially sampled every fifth patient (n=52) within each E/e′ tertile. Chart abstraction included key demographic variables, comorbid conditions, medication exposures (e.g., inotropes or pressors), laboratory values, echocardiography location (outpatient, non-ICU inpatient, ICU), and standard echocardiographic parameters (e.g., ejection fraction). Because initial sampling yielded a small number of grade 2 and grade 3 diastolic dysfunction cases, we enriched the sample by identifying 10 patients from the E/e′ >12 group and another 20 patients using pre-specified screening criteria of E/e′ >14 combined with E/A >2.^2^

All studies were performed on institutional clinical platforms by accredited sonographers using standard, ASE-recommended views. The following measurements were directly extracted from the archived images by a member of the study team blinded to LAP adjudication and ASE-assigned diastolic dysfunction grade: peak mitral inflow E-wave velocity, LVOT diameter, LVOT VTI and body surface area (BSA). Stroke volume was computed as the product of LVOT cross-sectional area and LVOT VTI, and stroke volume index (SVI) as stroke volume divided by BSA. Peak E-wave acceleration time (PEWAT, defined as the time interval from onset of the E-wave to its peak) was measured by study cardiologists (MM, AM), blinded to outcomes, from trans-mitral Doppler measurements (**Figure 1**). Standard measurements, including tissue Doppler e′ velocities (septal and lateral), tricuspid regurgitation velocity, and indexed left atrial volume, used for institutional ASE grading and LAP adjudication were adjudicated by cardiologist members of the study team (MM, AM).CEI was coded after completion of chart abstraction by the analyst (JD).

**Figure 1:**
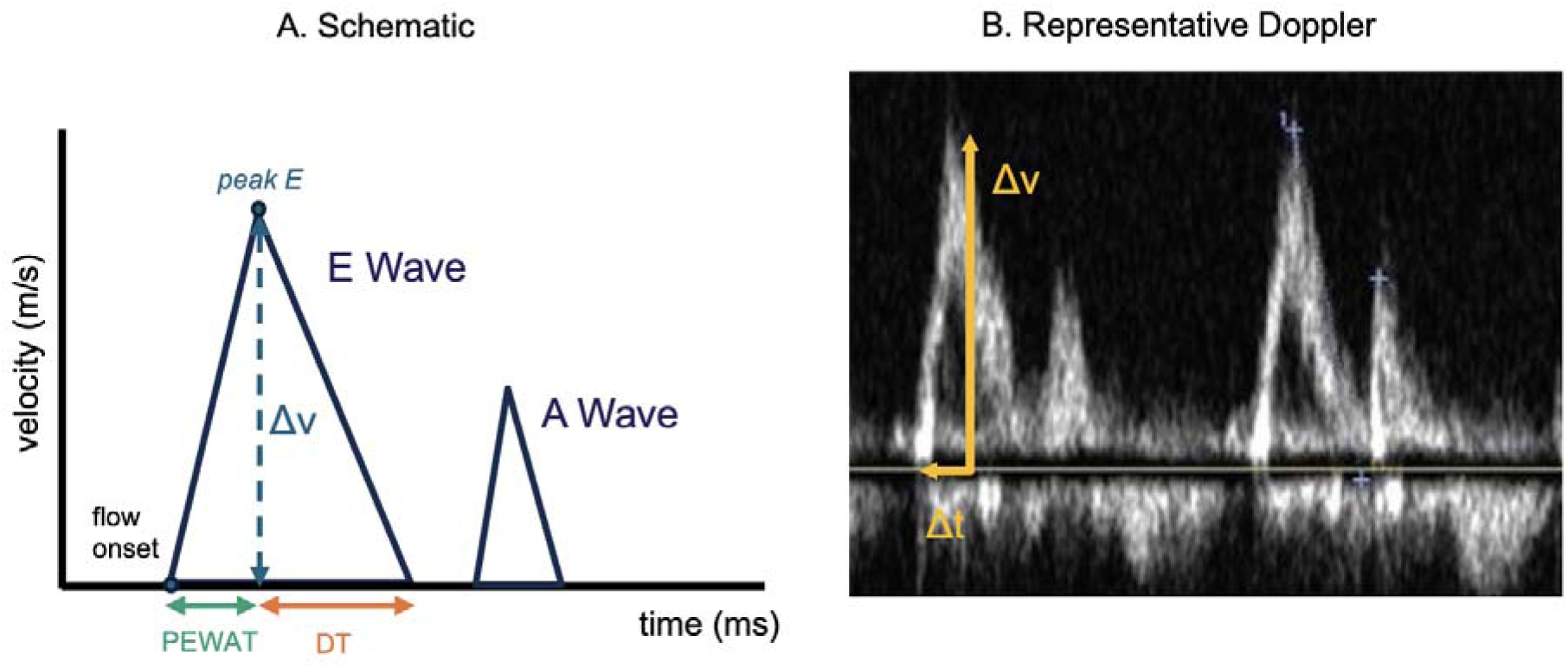
Measurement of Peak E-Wave Acceleration Time (PEWAT) from Trans-Mitral Doppler.

**Figure 2.**
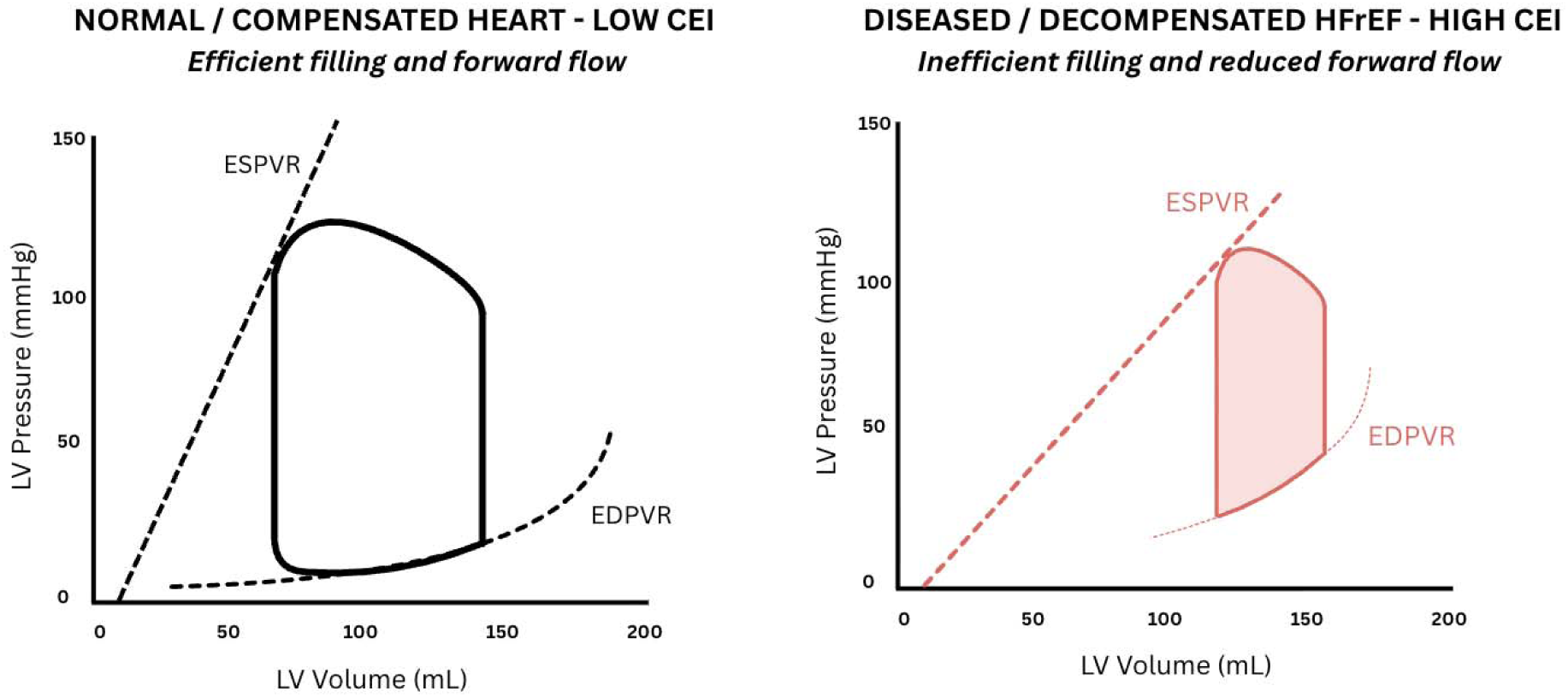
Cardiac Efficiency Index: Efficient Heart versus Inefficient Heart. Schematic LV pressure-volume loops illustrating efficient filling in a normal heart (left, low CEI) and inefficient filling in decompensated HFrEF (right, high CEI). Left: Steep ESPVR and relatively shallow EDPVR reflect preserved contractility and ventricular compliance, respectively, allowing the LV to accommodate diastolic filling with a limited rise in pressure while preserving stroke volume (SV). Right: Flatter ESPVR and steeper EDPVR reflect impaired contractility and reduced ventricular compliance, respectively, resulting in a greater rise in diastolic pressure and reduced SV. In this decompensated state, elevated filling pressures may promote a higher E-wave velocity and shorter PEWAT, reflecting more rapid development of the early transmitral pressure gradient and contributing, together with reduced SV, to a higher CEI. CEI, cardiac efficiency index; EDPVR, end-diastolic pressure-volume relationship; ESPVR, end-systolic pressure-volume relationship; HFrEF, heart failure with reduced ejection fraction; LV, left ventricle; PEWAT, peak E-wave acceleration time; SV, stroke volume.

ASE diastolic dysfunction grade was assigned according to the 2025 ASE multiparametric algorithm (grade 0 [normal], 1, 2, 3, or 4 [indeterminate]).¹ LAP was independently adjudicated as normal or elevated using ASE criteria.¹ Patients with grade 4 (indeterminate) diastolic dysfunction were excluded from analyses of diastolic dysfunction grade but retained in LAP analyses when LAP could be adjudicated.

#### Statistical analysis

Descriptive statistics were calculated for the overall LAP cohort and its constituent outcome subgroups (i.e., elevated vs. normal LAP), including assessment of missing data. Spearman rank correlations were used to evaluate associations between CEI and E/e′ and between CEI and each study outcome (LAP assessment and ASE diastolic dysfunction grade). Log transformation was planned for CEI if skewed. Sample size was determined by feasibility rather than a prespecified power calculation. A two-sided P<0.05 was considered statistically significant.

Binary discrimination for both outcomes was assessed with the area under the receiver-operating-characteristic curve (AUROC). Unadjusted logistic regressions were fit for E/e′ alone, CEI alone, and a combined model containing both. Ninety-five percent confidence intervals for AUROC were derived by nonparametric bootstrap resampling with 2,000 iterations.^14^ Threshold-specific operating characteristics (sensitivity, specificity, positive predictive value [PPV], negative predictive value [NPV]) were computed at the Youden-optimal threshold for each predictor against both the binary grade ≥2 outcome and the LAP (normal or elevated). Apparent point estimates were accompanied by Wilson 95% CIs, with bootstrap optimism correction subtracting mean optimism over 2,000 resamples. Stratum-specific odds ratios (ORs) across ordinal cut points were estimated by separate binary logistic regressions at each cut and are presented as exploratory due to small sample size. Ordinal discrimination across grade strata (collapsed as 0–1, 2, or 3) was assessed using proportional-odds logistic regression. Predictors were standardized to Z-scores so that odds ratios(ORs) express the change in odds of a higher-grade category per standard-deviation (SD) increase in the predictor. For each outcome, three models were fit: E/e′ alone, CEI alone, and combined. The proportional-odds assumption was assessed using the Brant test.^15^

Because the predictors operate on different scales and the upper grade strata were small, ordinal discrimination was summarized primarily with the c-index.^16^ As a sensitivity analysis addressing the reliability of PEWAT, the index was refit substituting deceleration time (DT), a routinely reported and well-validated measurement within diastolic dysfunction,^17^ in place of PEWAT.

All analyses were performed in R 4.5.^18^ This study followed the Strengthening the Reporting of Observational Studies in Epidemiology (STROBE) reporting guidelines.^19^

## Results

Of the 281 echocardiograms that were available, 82 were fully abstracted at convenience and met eligibility criteria with valid CEI measurements. Of these, 80 had a measurable LAP outcome and constituted the LAP analytic cohort; 66 were assigned an ASE grade of 0 (ie, normal), 1, 2, or 3 and constituted the Grade analytic cohort (**Supplementary Figure S1**). Fourteen patients with valid LAP estimates had an indeterminate ASE diastolic dysfunction grade, largely due to the presence of atrial fibrillation at the time of echocardiography.

Within the LAP cohort, median age was 74 years (interquartile range (IQR), 62–82), 46% were male, 81% had hypertension, 56% had ischemic heart disease, 39% had stage ≥3 chronic kidney disease, and 40% had a history of atrial fibrillation. Median left ventricular ejection fraction (LVEF) was 60% (IQR, 54–65), and median E/e′ was 13.0 (IQR, 10.0–16.4). NT-proBNP measured within 1 month before echocardiography was available in 49 patients, with a median value of 1,327 pg/mL (IQR, 369–5,275). Full baseline characteristics are summarized in **Table 1**. Elevated LAP was present in 33 of 80 patients (41.2%). Among patients with an assignable ASE diastolic dysfunction grade, 24 (36.4%) had grade 0, 20 (30.3%) grade 1, 11 (16.7%) grade 2, and 11 (16.7%) grade 3; overall, 22 (33.3%) had grade ≥2 diastolic dysfunction.

**Table 1.** Baseline clinical and echocardiographic characteristics, LAP cohort.

| **Characteristic** | **Normal LAP (n = 47)** | **Elevated LAP (n = 33)** | **Overall (n = 80)** |
|---|---|---|---|
| **Age, y, median (IQR)** | 73 (62–80) | 75 (62–83) | 74 (62–82) |
| **Male, n (%)** | 20 (43) | 17 (52) | 37 (46) |
| **Hypertension, n (%)** | 33 (70) | 32 (97) | 65 (81) |
| **Ischemic heart disease, n (%)** | 19 (40) | 26 (79) | 45 (56) |
| **CKD stage ≥ 3, n (%)** | 15 (32) | 16 (48) | 31 (39) |
| **Atrial fibrillation history, n (%)** | 9 (19) | 23 (70) | 32 (40) |
| **LVEF, %, median (IQR)** | 62 (58–66) | 55 (32–65) | 60 (54–65) |
| **E/e′ (average), median (IQR)** | 10.0 (8.3–12.8) | 17.6 (15.1–21.4) | 13.0 (10.0–16.4) |
| **CEI, median (IQR)** | 11 (8–16) | 38 (28–52) | 18 (10–33) |
| **NT-proBNP, pg/mL, median (IQR)** | 369 (132–834)<br>n = 17 | 2,466 (898–9,340)<br>n = 32 | 1,327 (369–5,275)<br>n = 49 |

*Continuous: median (IQR). Categorical: n (%). CKD = chronic kidney disease; CEI =Cardiac Efficiency Index; LAP = left atrial pressure; LVEF = left ventricular ejection fraction; NT-proBNP = N-terminal pro-B-type natriuretic peptide.*

CEI values were strongly right-skewed on the raw scale and were log-transformed for all regression analyses; the transformed distribution approximated normality (Shapiro-Wilk W = 0.98, P = 0.16). The log-transformed value was used throughout and is referenced as “CEI”. The distribution of CEI by adjudicated LAP status is shown in **Supplementary Figure S2**, with higher values in patients with elevated LAP (Mann-Whitney P < 0.001). CEI correlated strongly with E/e′ in the LAP cohort (Spearman ρ = 0.78, p <0.001; **Supplementary Figure S3**) and in the ASE Grade cohort (ρ = 0.76, p <0.001). The correlation between CEI and adjudicated LAP was ρ = 0.77 (p <0.001), and between CEI and ordinal ASE grade was ρ = 0.63 (p<0.001).

For the binary outcome of elevated LAP (n = 80; 33 cases, 47 controls), E/e′ yielded an AUROC of 0.949 (95% CI, 0.895–0.988) and CEI yielded an AUROC of 0.954 (95% CI, 0.898–0.993). A combined logistic model achieved an AUROC of 0.968 (95% CI, 0.922–0.999). For the binary outcome of ASE grade ≥ 2 versus grade 0–1 (n = 66; 22 cases, 44 controls), E/e′ yielded an AUROC of 0.942 (95% CI, 0.868–0.991) and CEI yielded an AUROC of 0.941 (95% CI, 0.854–0.994). A combined model achieved an AUROC of 0.958 (95% CI, 0.889–1.000). ROC curves are shown in **Figure 3**. At Youden-optimal thresholds (CEI ≥ 21 [log-CEI ≥ 3.05]; E/e′ ≥ 14), apparent sensitivity and specificity were 0.88 and 0.96 for CEI and 0.88 and 0.94 for E/e′ against elevated LAP; for grade ≥ 2 the corresponding values were 0.82 and 0.98 for CEI and 0.86 and 0.95 for E/e′. Bootstrap optimism correction reduced these estimates by 1-4%, consistent with stable threshold behavior under resampling (**Supplementary Table S2**). The six patients in whom CEI’s threshold disagreed with either adjudicated outcome are described at the patient level in **Supplementary Table S3**.

**Figure 3.**
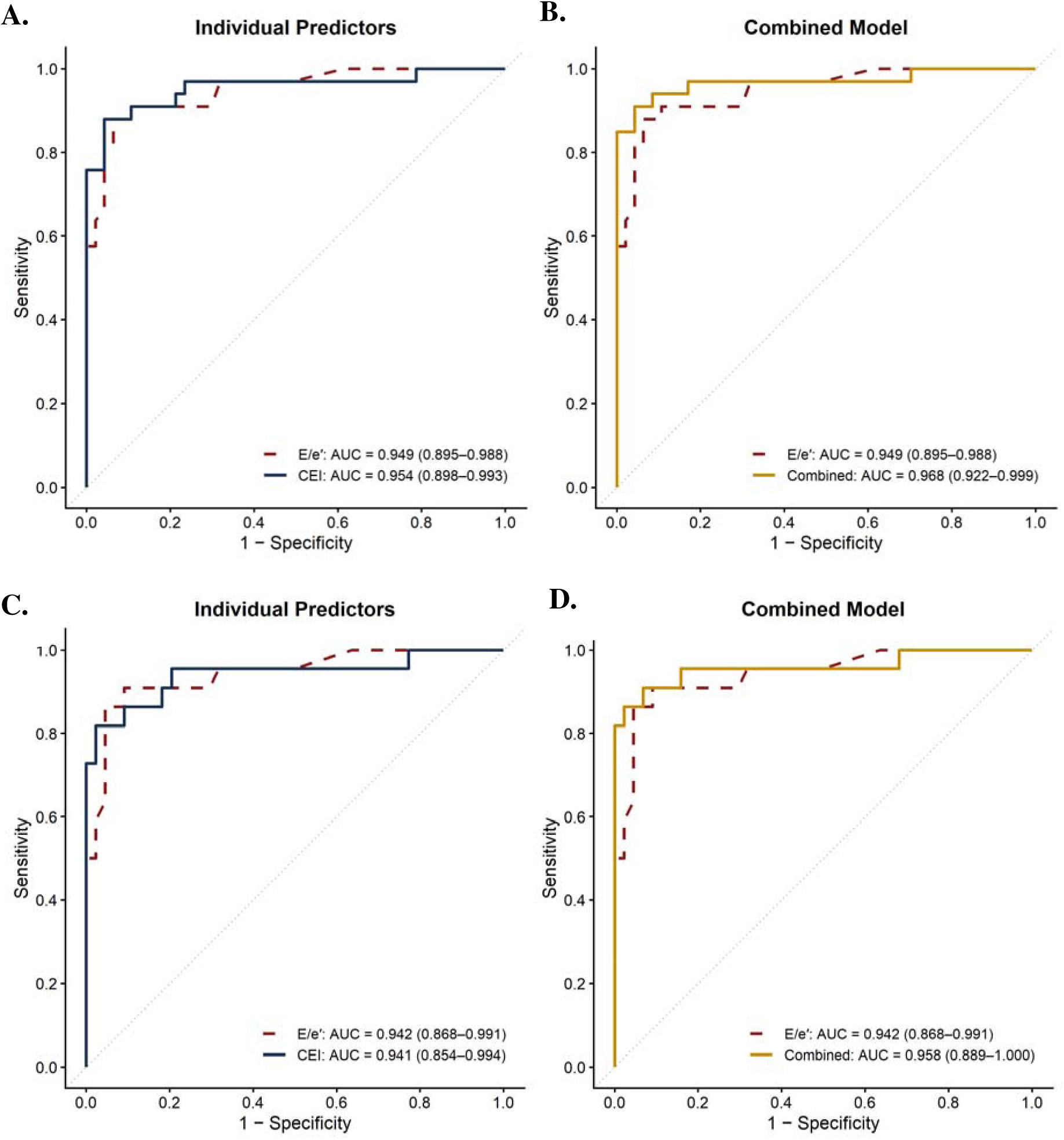
Receiver operating characteristic curves for elevated left atrial pressure and ASE grade ≥2 diastolic dysfunction. (A) Individual predictors and (B) combined model for elevated left atrial pressure (LAP; n=80; 33 cases, 47 controls). (C) Individual predictors and (D) combined model for ASE grade ≥2 diastolic dysfunction (n=66; 22 cases, 44 controls). AUCs are reported with bootstrapped 95% confidence intervals (2,000 iterations). ASE = American Society of Echocardiography; CEI = Cardiac Efficiency Index.

In ordinal logistic regression (grades 0–1, 2, 3; n = 66), CEI and E/e′ discriminated grade comparably (c-index 0.904 [95% CI, 0.830–0.968] and 0.912 [0.853–0.964], respectively), while CEI carried a larger per-SD association (OR 10.45 [95% CI, 4.03–27.08]; p <0.001) than E/e′ (OR 7.58 [2.92–19.67]; p <0.001) (**Supplementary Table S4**). In a combined model, CEI retained independent predictive value (OR 6.82 [2.24–20.73]; p <0.001) while E/e′ did not (OR 1.75 [0.70–4.38]; p=0.23); adding E/e′ to CEI did not improve fit (likelihood-ratio χ² = 1.87, p = 0.17), whereas adding CEI to E/e′ did (χ² = 14.21, p <0.001). The Brant test indicated violation of the proportional-odds assumption for both predictors (CEI p = 0.02; E/e′ p < 0.001), more pronounced for E/e′; ordinal discrimination is therefore summarized primarily by the c-index. In exploratory stratum-specific binary fits, the per-SD association was stronger at the grade 0–1 versus grade ≥ 2 cut than at the grade 0–2 versus grade 3 cut for both predictors (E/e′ approximately 138 and 4; CEI approximately 35 and 6). CEI followed a monotone pattern across ASE grade strata (**Figure 4**), with median values of 11 (Grade 0), 11 (Grade 1), 37 (Grade 2), and 52 (Grade 3).

**Figure 4.**
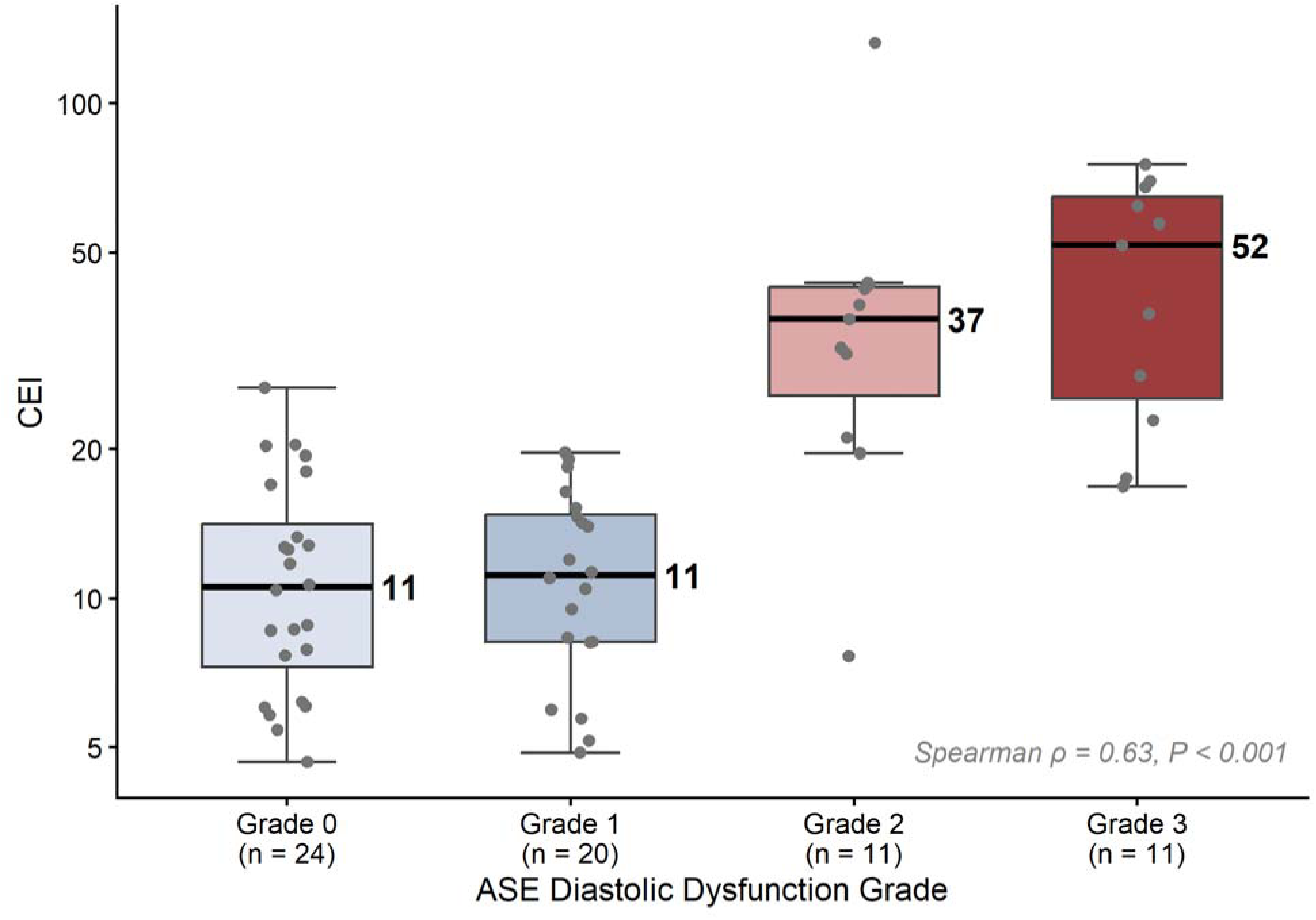
CEI by ASE Diastolic Dysfunction Grade. ASE Grade Cohort, n=66. Boxplots: median, IQR, Tukey whiskers; observations jittered. Spearman ρ *vs. ordinal grade = 0.63, P < 0.001*.

In the sensitivity analysis substituting deceleration time for PEWAT, AUROC was 0.919 for elevated LAP (n = 77) and 0.907 for ASE grade ≥ 2 (n = 65), with an ordinal c-index of 0.896.

## Discussion

In this retrospective proof-of-concept analysis, we introduce CEI, a novel and intuitive echocardiographic parameter that integrates early diastolic filling dynamics with forward SV, and provide preliminary evidence supporting its potential utility in the noninvasive assessment of diastolic dysfunction. CEI demonstrated strong discrimination of elevated LAP and ASE diastolic dysfunction grade ≥2, with performance comparable to E/e′ despite the inherent advantage of E/e′ being incorporated into the ASE criteria used to define both reference outcomes.

Unlike conventional diastolic parameters, CEI does not characterize ventricular filling in isolation but instead relates early diastolic inflow kinetics to the forward SV generated. This relationship provides the conceptual basis for CEI as a measure of cardiac efficiency: a higher CEI reflects greater early diastolic pressure-gradient development relative to forward SV and therefore less efficient physiology, whereas a lower CEI reflects greater forward SV relative to early diastolic pressure-gradient development and more efficient physiology. Consistent with this framework, higher CEI values were associated with elevated LAP and greater ASE diastolic dysfunction grade, whereas lower values were associated with lower LAP and preserved diastolic function.

If validated against invasive hemodynamics, CEI may provide a complementary approach to existing echocardiographic parameters for assessing diastolic physiology and estimating filling pressures.

Importantly, CEI is derived entirely from routinely acquired echocardiographic measurements and therefore requires no additional image acquisition. These measurements are also amenable to automated extraction and calculation, creating the potential for CEI to be integrated into echocardiographic analysis software and generated automatically as part of routine clinical interpretation.

### Limitations

This study has several limitations. First, the retrospective design and small convenience sample limit generalizability. Second, CEI was not validated against invasive hemodynamic measurements. Consequently, in cases where CEI and ASE classification were discordant, it remains unclear whether CEI reflected misclassification or differences in the underlying hemodynamic state. Third, CEI was assessed at a single time point and was not evaluated longitudinally within individual patients. Further investigation is therefore needed to determine whether serial changes in CEI track changes in ASE diastolic dysfunction grade and, more importantly, invasively measured filling pressures over time. Fourth, enrichment for patients with higher E/e′ and more advanced diastolic dysfunction may have increased discrimination performance relative to an unselected clinical population. Although this enrichment facilitated proof-of-concept evaluation across a broader spectrum of diastolic dysfunction, performance will require confirmation in unselected populations. Fifth, interobserver reproducibility of manual PEWAT measurements was not formally assessed and should be evaluated in future studies. Finally, external validation in independent cohorts using robust reference standards, particularly invasive hemodynamics, is essential before clinical application.

### Conclusion

CEI is a novel echocardiographic parameter that integrates early diastolic inflow kinetics with forward stroke volume within a conceptual framework of cardiac efficiency. In this proof-of-concept analysis, CEI demonstrated strong discrimination of elevated LAP and ASE diastolic dysfunction grade, with higher CEI values associated with greater diastolic dysfunction severity. These findings support further investigation and prospective validation of CEI against invasive hemodynamics in larger, more generalizable cohorts.

## Disclosures

Davis is an ad-hoc consultant for GE Healthcare on ultrasound and point-of-care ultrasound related innovations, but has not received payment for any consulting in the last 18 months. May is an ad-hoc consultant for GE Healthcare for electrocardiogram-related innovations unrelated to the present study. Davis and May hold intellectual property (in conjunction with their affiliated institution) that covers the commercialization of this parameter. It is currently unlicensed. All authors were blinded to at least one critical component of the cohort data (echocardiography raw data, statistical analysis, or both) to avoid the appearance of conflicts of interest influencing results. De-identified data and scripts for statistical analyses are available on request.

The remaining authors have nothing to disclose.

## Data Availability

All data produced in the present study are available upon reasonable request to the authors

## Supplementary Materials

**Supplementary Figure S1.**
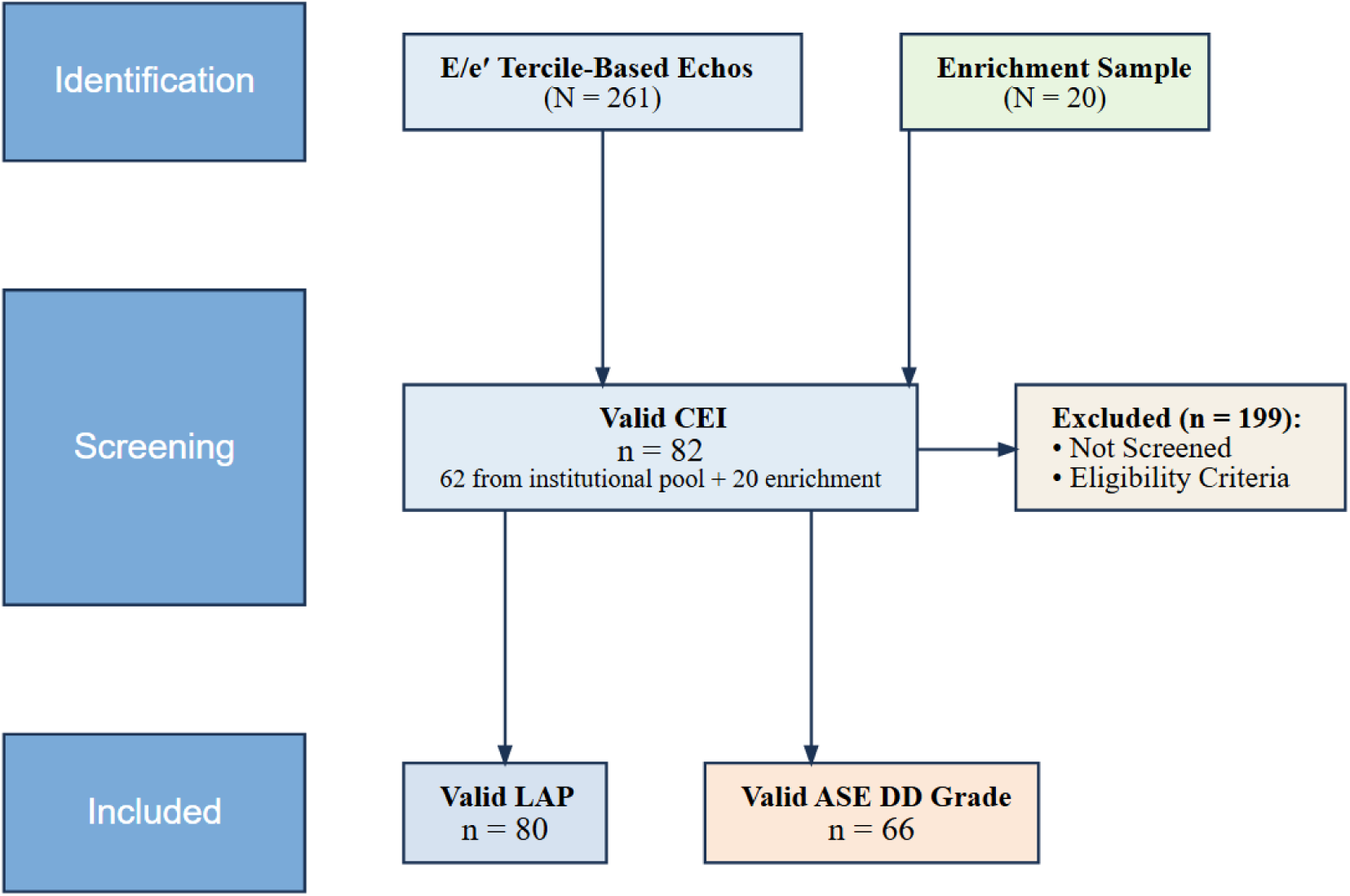
Cohort derivation. Both cohorts derive from the same CEI-valid sample (n = 82). 14 patients with grade 4/indeterminate ASE assignment had measurable LAP and contribute only to the LAP cohort. CEI = Cardiac Efficiency Index; LAP = left atrial pressure.

**Supplementary Figure S2.**
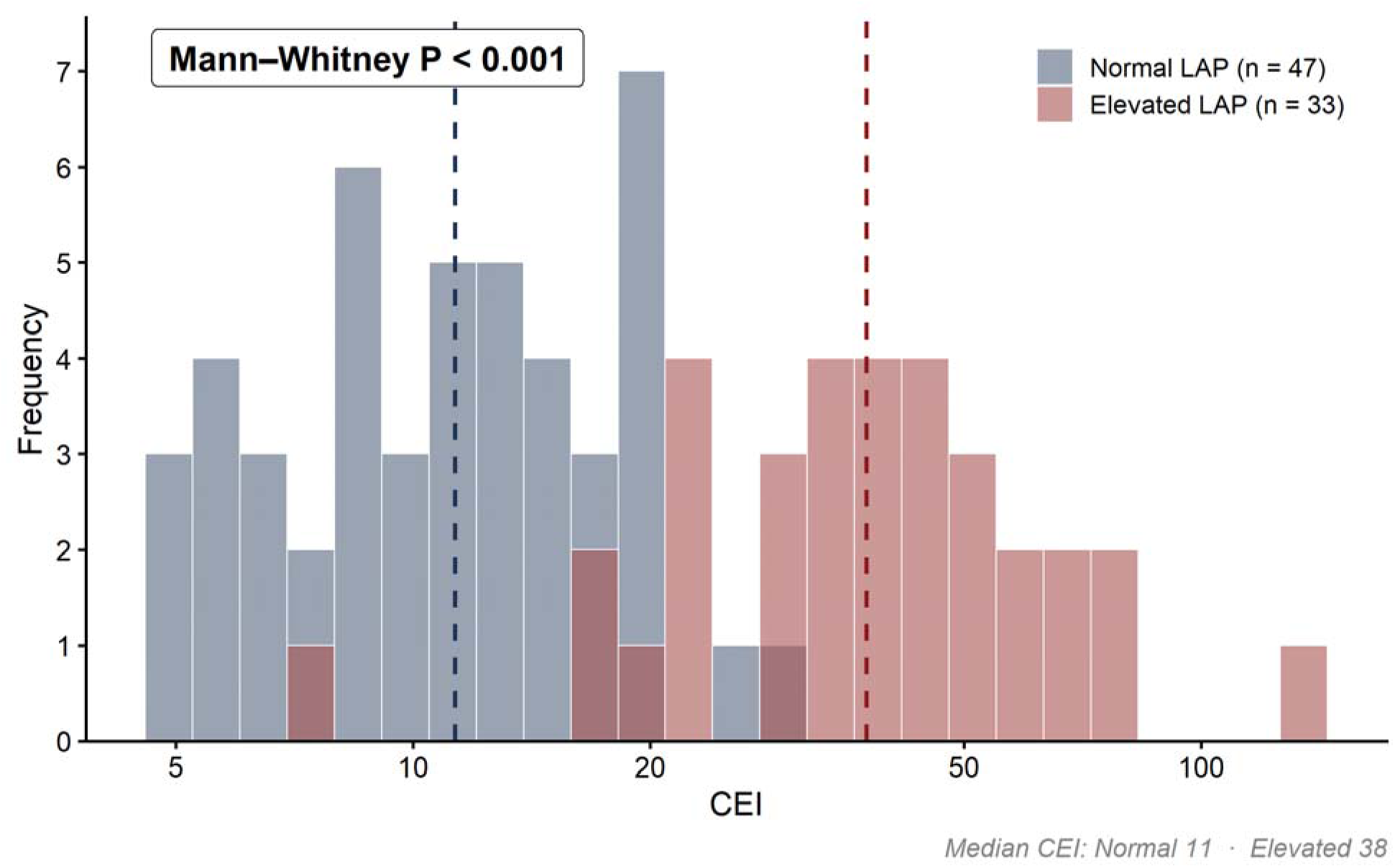
Distribution of CEI by LAP. Dashed lines = group medians. Mann-Whitney P < 0.001.

**Supplementary Figure S3.**
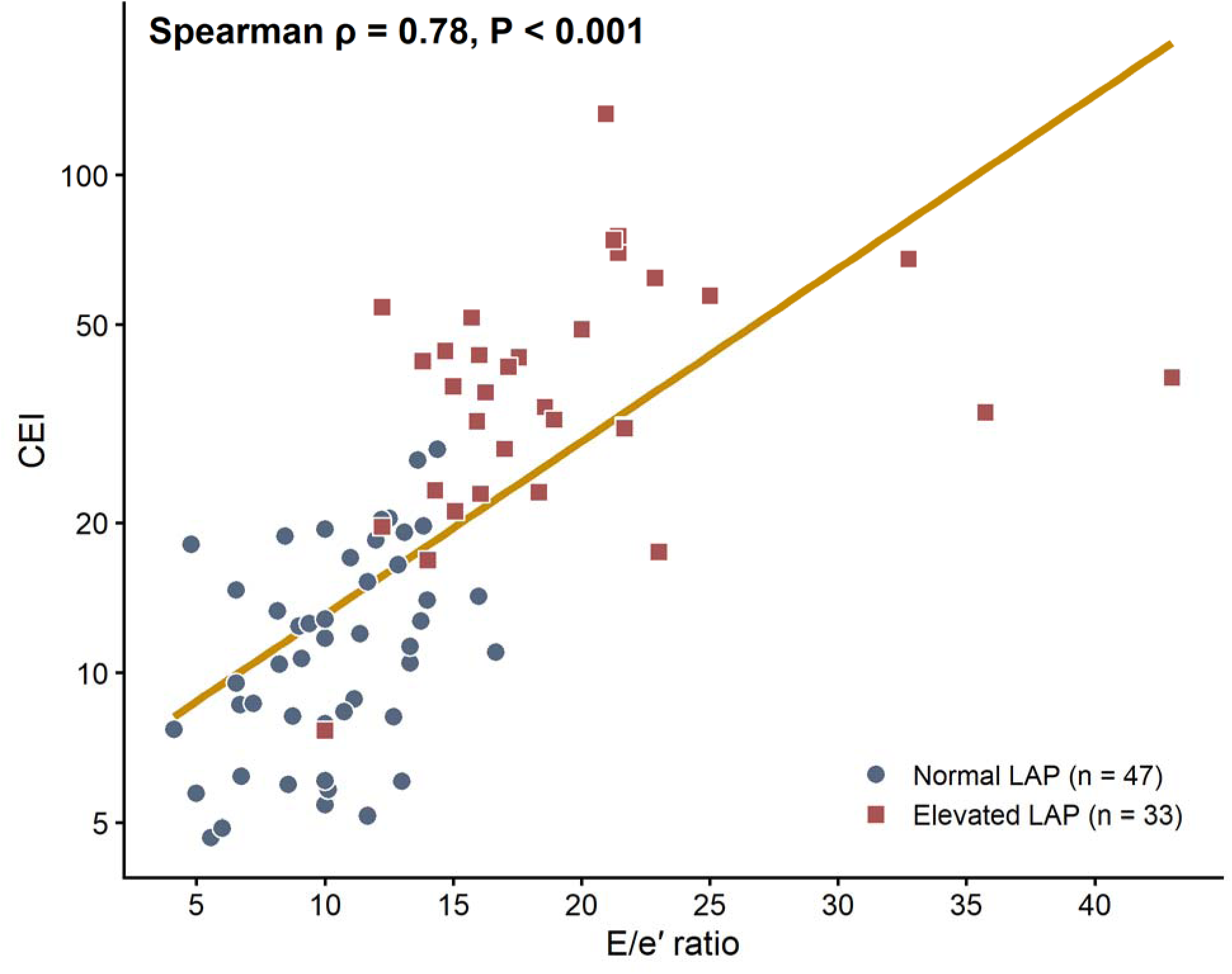
Correlation of CEI with E/e′, LAP cohort. LAP Cohort, n = 80: Spearman ρ = 0.78, P < 0.001. ASE Grade Cohort, n = 66: ρ = 0.76.

**Supplementary Table S1.** Interpretation of CEI in Different Clinical States.

| **Expected CEI** | **Representative Clinical State** | **Diastolic ΔP/Δt (4E<sup>2</sup>/PEWAT)** | **Stroke Volume Index** | **Physiologic Interpretation** |
|---|---|---|---|---|
| **Low to intermediate** | **Normal to near-normal physiology** — Normal LV geometry; normal or impaired-relaxation mitral inflow pattern; normal filling pressures; preserved cardiac output | **Low to intermediate** | **Preserved** | Relatively modest early diastolic pressure-gradient development with preserved forward flow, consistent with efficient translation of ventricular filling into forward output. |
| **Intermediate to high** | **HFpEF** — Normal LV size or concentric remodeling/hypertrophy; pseudonormal or restrictive mitral inflow; elevated filling pressures; preserved LVEF | **High** | **Preserved to mildly reduced** | Increased early diastolic pressure-gradient development relative to forward flow, resulting in an intermediate-to-high CEI despite preserved LVEF. |
| **High** | **Advanced HFrEF / cardiogenic shock** — LV systolic dysfunction; restrictive mitral inflow; markedly elevated filling pressures; reduced cardiac output | **High** | **Low** | Marked early diastolic pressure-gradient development with reduced forward flow, consistent with inefficient translation of ventricular filling into forward output. |
| **High** | **Decompensated severe mitral regurgitation** — Prominent early mitral inflow; elevated filling pressures; reduced effective forward output | **High** | **Low effective forward SVI** | Marked early diastolic pressure-gradient development with reduced effective forward stroke volume, resulting in a high CEI despite potentially preserved total LV stroke volume. |
| **Intermediate** | **Severely reduced cardiac preload** — Reduced early mitral inflow; low filling pressures; reduced cardiac output | **Low** | **Low** | Low early diastolic pressure-gradient development accompanied by reduced forward flow, distinguishing reduced ventricular filling from disproportionate inflow kinetics relative to forward output. |

*Representative physiologic states illustrate the relationship between early diastolic pressure-gradient development, forward stroke volume, and expected Cardiac Efficiency Index. CEI reflects the relationship between early mitral inflow kinetics and forward SVI: lower values indicate greater forward SVI relative to early inflow kinetics, whereas higher values indicate greater early inflow kinetics relative to forward SVI. These clinical states are illustrative and are not intended to represent diagnostic criteria or established CEI thresholds.*
***Abbreviations:*** *CEI, Cardiac Efficiency Index; HFpEF, heart failure with preserved ejection fraction; HFrEF, heart failure with reduced ejection fraction; LV, left ventricle; LVEF, left ventricular ejection fraction; PEWAT, peak E-wave acceleration time; SVI, stroke volume index; ΔP/Δt, rate of LA-to-LV pressure-gradient development.*

**Supplementary Table S2.** Threshold-specific operating characteristics for CEI and E/e ′ against elevated left atrial pressure and ASE diastolic dysfunction grade ≥ 2.

| **Predictor / Statistic** | **LAP: Apparent (95% CI)** | **LAP: Optimism-corrected (95% CI)** | **ASE Grade: Apparent (95% CI)** | **ASE Grade: Optimism-corrected (95% CI)** |
|---|---|---|---|---|
| ***CEI (≥3.05)*** |  |  |  |  |
| **Sensitivity** | 0.88 (0.73–0.95) | 0.85 (0.69–0.94) | 0.82 (0.61–0.93) | 0.78 (0.57–0.90) |
| **Specificity** | 0.96 (0.86–0.99) | 0.94 (0.84–0.98) | 0.98 (0.88–1.00) | 0.96 (0.86–0.99) |
| **PPV** | 0.94 (0.79–0.98) | 0.92 (0.77–0.97) | 0.95 (0.75–0.99) | 0.92 (0.72–0.98) |
| **NPV** | 0.92 (0.81–0.97) | 0.90 (0.79–0.96) | 0.91 (0.80–0.97) | 0.90 (0.78–0.96) |
| ***E/e' (≥14)*** |  |  |  |  |
| **Sensitivity** | 0.88 (0.73–0.95) | 0.86 (0.70–0.94) | 0.86 (0.67–0.95) | 0.83 (0.63–0.94) |
| **Specificity** | 0.94 (0.83–0.98) | 0.93 (0.82–0.97) | 0.95 (0.85–0.99) | 0.95 (0.84–0.98) |
| **PPV** | 0.91 (0.76–0.97) | 0.89 (0.74–0.96) | 0.90 (0.71–0.97) | 0.89 (0.69–0.97) |
| **NPV** | 0.92 (0.80–0.97) | 0.90 (0.79–0.96) | 0.93 (0.82–0.98) | 0.92 (0.80–0.97) |

*Apparent estimates are the in-sample point estimates at the full-sample Youden-optimal threshold, with Wilson 95% CIs. Optimism-corrected estimates subtract mean optimism over 2,000 bootstrap resamples. The Youden-optimal threshold for CEI was identical (log-CEI ≥ 3.05) for both outcomes; for E/e' the threshold was ≥ 14 for both outcomes. Because the threshold is derived in-sample, apparent values are subject to optimism bias and require external validation. ASE = American Society of Echocardiography; CEI = Cardiac Efficiency Index; LAP = left atrial pressure; NPV = negative predictive value; PPV = positive predictive value.*

**Supplementary Table S3.** CEI misclassifications at the Youden-optimal threshold across the LAP and ASE grade ≥ 2 outcomes.

| **Case** | **LVEF** | **E** | **PEWAT** | **SVI** | **E/e′** | **CEI** | **Grade** | **LAP** | **AF status** | **Discordant Outcome** |
|---|---|---|---|---|---|---|---|---|---|---|
| **1** | 68 | 90 | 65 | 65.2 | 10.0 | 8 | 2 | Elev. | — | LAP + Grade ≥ 2 |
| **2** | 64 | 95 | 65 | 33.0 | 14.0 | 17 | 3 | Elev. | — | LAP + Grade ≥ 2 |
| **3** | 36 | 109 | 97 | 28.0 | 23.0 | 17 | 3 | Elev. | Hx AF | LAP + Grade ≥ 2 |
| **4** | 65 | 110 | 56 | 44.0 | 12.2 | 20 | 2 | Elev. | Hx AF | LAP + Grade ≥ 2 |
| **5** | 64 | 95 | 43 | 31.4 | 13.6 | 27 | 0 | Norm. | Hx AF | LAP + Grade ≥ 2 |
| **6** | 68 | 94 | 63 | 20.0 | 14.4 | 28 | — | Norm. | Hx AF | LAP |

*AF = atrial fibrillation; PEWAT =* peak E-wave acceleration time *(ms); E = peak E-wave velocity (cm/s); Elev. = elevated; Hx = history of; LVEF = left ventricular ejection fraction (%); Norm. = normal; SVI = indexed stroke volume (mL/m<sup>2</sup>).*

**Supplementary Table S4.** Ordinal logistic regression, ASE Grade (0-1, 2, 3).

| **Model** | **OR per SD (95% CI)** | **P value** | **c-index** | **Brant P** |
|---|---|---|---|---|
| **E/e′ alone** | 7.58 (2.92–19.67) | < 0.001 | 0.912 | <0.001 |
| ***E/e': grade 0–1 vs ≥ 2*** | 138.12 (9.07–2103.61) | < 0.001 | — | — |
| ***E/e': grade 0–2 vs 3*** | 3.93 (1.57–9.81) | 0.003 | — | — |
| **CEI alone** | 10.45 (4.03–27.08) | < 0.001 | 0.904 | 0.02 |
| ***CEI: grade 0–1 vs ≥ 2*** | 35.43 (5.46–229.92) | < 0.001 | — | — |
| ***CEI: grade 0–2 vs 3*** | 5.65 (2.13–14.98) | < 0.001 | — | — |
| **Combined: CEI + E/e′** | 6.82 (2.24–20.73) - CEI<br>1.75 (0.70–4.38) - E/e′ | < 0.001<br>0.23 | 0.908 | 0.02 |

*OR per SD increase in predictor. c-index is model concordance (Somers’ D rank-correlation between predicted and observed grade). Brant P < 0.05 indicates violation of the proportional-odds assumption. ASE = American Society of Echocardiography; CI = confidence interval; DD = diastolic dysfunction; CEI = Cardiac Efficiency Index; OR = odds ratio; SD = standard deviation.*

## Notes

### Author Declarations

The Washington University's Institutional Review Board approved the study and its design.

